# Lesioned hemisphere-specific phenotypes of post-stroke fatigue emerge from motor and mood characteristics in chronic stroke

**DOI:** 10.1101/2023.09.01.23294971

**Authors:** William De Doncker, Annapoorna Kuppuswamy

## Abstract

Post-stroke fatigue commonly presents alongside several co-morbidities. The interaction between co-morbidities and their relationship to fatigue is not known. Here we focus on physical and mood comorbidities, alongside lesion characteristics. We predict the emergence of distinct fatigue phenotypes with distinguishable physical and mood characteristics.

In this cross-sectional observational study, in 94 first time, non-depressed, moderate to minimally impaired chronic stroke survivors, the relationship between measures of motor function (Grip strength, Nine Hole Peg Test time), motor cortical excitability (Resting Motor Threshold), Hospital Anxiety and Depression Scale scores and Fatigue Severity Scale – 7 scores age, gender and side of stroke was established using Spearman’s rank correlation. Mood and motor variables were then entered into k-means clustering algorithm to identify number of unique clusters, if any. Post-hoc pairwise comparisons followed by corrections for multiple comparisons was performed to characterise differences between clusters in the variables included in k-means clustering.

Clustering analysis revealed a 4-cluster model to be the winning model (average silhouette score of 0.311). There was no significant difference in FSS-7 scores between the 4 high-fatigue clusters. 2 clusters consisted of only left hemispheric strokes, and the remaining two were exclusively right hemisphere strokes. Factors that differentiated hemisphere-specific clusters was the level of depressive symptoms and anxiety. Motor characteristics distinguished the low-depressive left from right hemispheric clusters.

The significant differences in side of stroke, and the differential relationship between mood and motor function in the 4 clusters reveals the heterogenous nature of post-stroke fatigue, which is amenable to categorisation. Such categorisation is critical for understanding the interactions between post-stroke fatigue and its presenting comorbid deficits, with significant implications for development of context/category specific interventions.

## Introduction

Post-stroke fatigue is a significant and highly prevalent symptom in chronic stroke^1–3^. Although commonly seen alongside other post-stroke disturbances such as depression, anxiety, cognitive dysfunction and motor impairment, fatigue is an independent symptom and is poorly understood^4–8^.

An extensive body of literature is dedicated to studying the relationship between fatigue, lesion location, impairment and mood disorders^6,9–13^. While fatigue incidence is greater when lesion includes the thalamus, when there is significant depression and anxiety, and sometimes, in the presence of physical and cognitive impairment, it is still unclear how these different factors interact and, in combination, explain fatigue incidence and severity. For example, in some chronic stroke survivors, fatigue can emerge in the absence of physical, cognitive or mood deficits, while in others it occurs in the presence of one or more deficits. It is critical we identify the contexts i.e. combination of factors that co-present with fatigue, and establish if there are certain contexts more conducive for fatigue. The need for such understanding arises from intervention studies which show that responsiveness to a treatment is dependent on the context (phenotype) of fatigue^14^. In this study we aim to identify fatigue phenotypes, based on factors related to post-stroke fatigue.

Low motor cortical excitability of the stroke lesioned hemisphere is seen in high post-stroke fatigue^15^, and will be included as the neural correlate of fatigue for phenotyping. While side of lesion is not associated with either incidence or severity of fatigue, balance of interhemispheric inhibition is reversed, with a right hemispheric dominance associated with high fatigue^16^. This shift in interhemispheric inhibition is independent of side of stroke indicating that shift in laterality is not related to stroke lesion, but fatigue. The impact of shift in laterality of inhibition, on motor cortical excitability of the lesioned hemisphere is not known, but is likely to be influenced. Therefore, side of stroke lesion, motor cortical excitability, along with measures of hand motor function will be included as physical factors in phenotyping fatigue.

Post-stroke fatigue is correlated with depression and anxiety^10^; and measures of mood will be included in phenotyping fatigue. There is a known effect of stroke side on depression^17^, with left hemisphere strokes carrying a greater burden of depression. Given the impact of side of stroke on both motor physiology and mood, to avoid large side-related effects on depression overshadowing its effect on motor physiology, we exclude those with clinically diagnosed depression.

To summarise, in this investigation, we first identify the association between post-stroke fatigue, motor function, motor physiology, depressive symptoms and anxiety, followed by categorisation of high fatigue into clusters, to define the motor and mood characteristics of each category. We expect to replicate previous associations of post-stroke fatigue, with an emergence of distinguishable clusters driven by motor, mood and side of stroke.

## Materials and Methods

### Subjects

This cross-sectional observational study was approved by the London Bromley Research Ethics Committee (REC reference number: 16/LO/0714). 94 stroke survivors who were (1) first-time stroke; (2) ≥ 3 months post-stroke were included in the study. Exclusion criteria were (1) diagnosis of any other neurological disorder; (2) on anti-depressants or centrally acting drugs. (3) depression scores ≥ 11 assessed using Hospital Anxiety and Depression Scale (HADS); (4) grip strength or manual dexterity (Nine-hole-peg-test) of the affected hand <60% of unaffected hand; (5) contraindications to Transcranial Magnetic Stimulation (Table 1: demographics of the 94 stroke survivors). All stroke survivors provided written informed consent in accordance with the Declaration of Helsinki.

**Table 1.** Demographics of patients that completed the study. Mean (SD) values are displayed for all continuous variables and number of patients are displayed for categorical variables (Hemisphere affected, gender, dominant hand). P-value indicate the significance of spearman rank correlations between FSS-7 and continuous variables and Wilcoxon rank sum tests comparing the FSS-7 score within categorical values for the entire patient cohort and the two groups divided based on hemisphere affected by the stroke.

|  | All patients (N = 98) | Left Hemisphere Stroke (N = 51) | Right Hemisphere Stroke (N = 47) | P-value |
| --- | --- | --- | --- | --- |
| **FSS-7** | 3.83 (1.91) | 3.59 (1.94) | 4.09 (1.85) |  |
| Mean (SD) |  |  |  |  |
| **Hemisphere Affected** |  |  |  | *P* = 0.20 |
| Left \| Right | 51 \| 47 |  |  |  |
| **Gender** |  |  |  | *P* = 0.003; 0.002; 0.200 |
| Females \| Males | 32 \| 66 | 21 \| 30 | 11 \| 36 |  |
| **Dominant Hand** |  |  |  | *P* = 0.128; 0.765; 0.064 |
| Right \| Left | 88 \| 10 | 47 \| 4 | 41 \| 6 |  |
| **Age**<sub>years</sub> | 60.93 (12.65) | 59.69 (13.40) | 62.28 (11.78) | *P* = 0.050; 0.008; 0.911 |
| **Grip**<sub>% unaffected hand</sub> | 91.37 (22.93) | 96.96 (19.14) | 85.31 (25.27) | *P* = 0.044; 0.454; 0.130 |
| **NHPT**<sub>% unaffected hand</sub> | 88.48 (24.63) | 97.98 (22.46) | 78.16 (22.85) | *P* = 0.226; 0.689; 0.212 |
| **HADS**<sub>Depression</sub> | 4.83 (3.23) | 4.29 (3.11) | 5.40 (3.30) | *P* < 0.001; < 0.001; 0.002 |
| **HADS**<sub>Anxiety</sub> | 5.26 (3.93) | 4.71 (4.25) | 5.85 (3.49) | *P* < 0.001; 0.0084; 0.0162 |
| **RMT**<sub>Affected Hemisphere (% MSO)</sub> | 47.43 (11.90) | 48.47 (11.88) | 46.30 (11.94) | *P* = 0.791; 0.210; 0.416 |

### Fatigue quantification

Fatigue was measured using the Fatigue Severity Scale (FSS-7), a widely used and validated questionnaire to measure self-reported fatigue across different conditions including stroke^18^. An average score of 4 out of a maximum of 7 was used as the lower limit for defining ‘high fatigue’ group, as commonly used in clinical practice.

### Mood quantification

Hospital Anxiety and Depression Questionnaire, a validated widely used questionnaire was used to measure both anxiety and depressive symptoms. A maximum score of 21 can be obtained each for the Anxiety and Depression sub-sections. A score of 11 or below is considered to be in the non-depressive range, although higher the number, greater the likelihood of depression like symptoms. While we excluded anyone with greater than 11 in the depression sub-section score, there was no exclusion based on anxiety scores.

### Motor function

Hand grip strength was measured using a hand-held dynamometer. Three repetitions of maximum voluntary grip were performed on each hand in a randomised order with adequate rest between contractions, with the average of 3 contractions taken as the Maximum Voluntary Force for a given hand. Grip function was calculated as follows

Grip = average grip [(affected hand)/ average grip (unaffected hand)] x 100

The average time taken to complete three repetitions of a nine-hole peg test was taken as dexterity score. Dexterity function was calculated as follows

Dexterity = average dexterity score [(affected hand)/ average dexterity score (unaffected hand)] x 100

Motor physiology - Surface electromyogram and transcranial magnetic stimulation

Electromyogram (EMG) recordings were obtained from the FDI muscle using surface neonatal prewired disposable electrodes (1041PTS Neonatal Electrode, Kendell) in a belly-tendon montage with the ground positioned over the flexor retinaculum of the hand. The signal was amplified with a gain of 1000 (D360, Digitmer, Welwyn Garden City, UK), bandpass filtered (100-1000 Hz), digitized at 5kHz (Power1401, CED, Cambridge, UK) and recorded with Signal version 6.04 software (CED, Cambridge, UK).

A TMS device (Magstim 200^2^, Magstim, Whitland, Wales) connected to a figure-of-eight coil (wing diameter, 70mm) was used to stimulate the hand area of M1 in each hemisphere. The coil was held tangentially on the scalp at an angle of 45° to the mid-sagittal plane to induce a posterior-anterior (PA) current across the central sulcus. The subjects were instructed to stay relaxed with their eyes open and their legs uncrossed. The motor ‘hotspot’ of the FDI muscle for each hemisphere was determined as previously^15^.

Resting motor threshold (RMT) for each hemisphere was defined as the lowest intensity of stimulation (% MSO) required to evoke a peak-to-peak MEP amplitude at the hotspot of at least 50μV in a minimum of 5 of 10 consecutive trials while subjects were at rest.

### Statistical Analysis

Spearman’s Rank Correlations between FSS-7 and age, anxiety, depression, grip strength, NHPT, SDMT and RMT were calculated. Wilcoxon rank sum tests were used to assess the difference in FSS-7 across different groups divided based on gender, hemisphere affected and dominant hand being affected.

A multiple linear regression analysis was used to examine the effect of RMT on FSS-7 in all patients and in two groups based on the hemisphere affected by the stroke. Assumptions of normality and homoscedasticity of the residuals for each linear regression model were assessed visually using quantile-quantile normal plots and fitted-versus residual-value plots. The level of significance was set at p = 0.05.

#### k-means clustering analysis

Of the 94 patients included in the regression analyses, 47 had an FSS-7 score of ≥ 4. This sub-set of 47 was defined as the high fatigue group and were subject to k-means clustering analysis. Motor variables of grip, NHPT and RMT were compressed into a single variable using Principal Component Analysis. The average of the first 2 principal components was used as the motor PCA score. The first component almost exclusively represented data from functional measures and second principal component represented data from RMT, hence the average of the first 2 components to represent the overall motor score. Z-scores were calculated for the 4 variables of interest (HADS-Anxiety, HADS-depression, stroke side and motor PCA) and were entered into the k-means clustering algorithm. A k-means cluster analysis is an unsupervised machine learning algorithm-based technique used to identify subgroups (or clusters) in a dataset that represent data points that are similar to one another, yet distinct from data points in other clusters. The *k*-means algorithm clusters the data into a number, *k*, of predefined, distinct, and non-overlapping groups where each data point only belongs to one group. Data points are assigned to a particular cluster in such a way that the sum of the squared distance between all of the data points, and the mean of all the data points that belong to that cluster, is minimized. Based on known associations between post-stroke fatigue, motor function, motor physiology and mood characteristics, we tested k’s of 2-5. The best model was chosen from Silhouette scores, a score that indicates robustness of clustering. A score of 1 indicates completely non-overlapping clusters, while a score of 0 means there is no difference between clusters, a score of -1 indicates wrong classification. A score between 0.3 and 0.7 is normally considered acceptable for identifying unique non-overlapping clusters. The model with the highest silhouette score above 0.3 was considered the winning model in this study, and the number of clusters determined from this model. Given the non-parametric nature of the data, a Kruskal-Wallis Analysis of Variance test, followed by post-doc comparisons using Dunn’s method adjusted for multiple comparisons using Bonferroni correction, were performed to identify differences between the various clusters in the parameters of interest.

## Results

There was a significant association between FSS-7 and grip strength (rho=-0.204, p=0.044), HADS_Anxiety_ (rho=0.37, p<0.001) and HADS_Depression_ (rho=0.50, p<0.001) in 94 participants. When patients were divided into groups based on hemisphere affected, there was a significant association between FSS-7 and age (rho=-0.37, p=0.008), anxiety (rho=0.37, p=0.008) and depression (rho=0.53, p<0.001) in left hemisphere strokes and a significant positive association between FSS-7 and HADS_Anxiety_ (rho=0.35, p=0.016) and HADS_Depression_ (rho=0.44, p=0.002) in right hemisphere strokes. There was a significant difference in median FSS-7 score between males and females in the entire cohort and left hemisphere strokes (p=0.003; p=0.002). There was no association between FSS-7, ARAT and RMT affected hemisphere and no difference in FSS-7 in left hemisphere strokes compared to right hemisphere strokes. Patient demographics can be found in table 1.

The best fit regression model that predicted FSS-7 in the entire sample included RMT, HADS – A and Age (F_(3,94)_=5.97, p<0.001), with an adjusted R^2^ of 0.133 (figure 1A). When patients were divided into two groups based on stroke hemisphere, in left hemisphere stroke,, the combined predictive power of RMT (beta=0.047, p=0.025, CI[0.006,0.090]), HADS_Anxiety_ (beta=0.162, p=0.008, CI[0.044,0.280]) and age (beta=-0.044, p=0.017, CI[-0.081,-0.008]) explained FSS-7 scores (F_(3,47)_=6.38, p=0.001), with an adjusted R^2^ of 0.244 (figure 1C).. There was no significant FSS-7 predictors for right hemisphere strokes (F_(3,43)_=2.16, p=0.106) (figure 1B).

**Figure 1.**
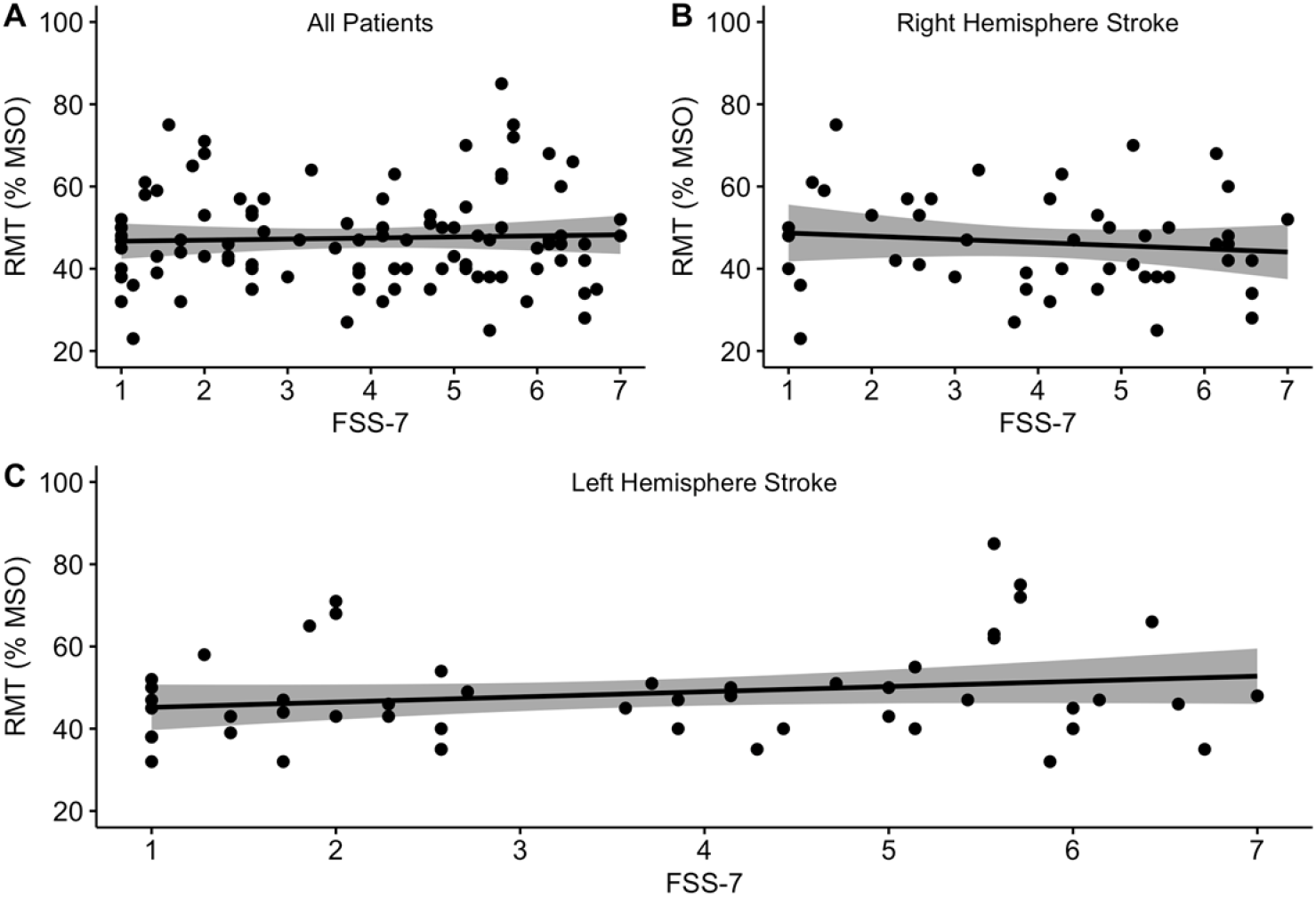
Regression line with 95% confidence interval and individual data points displaying the association between resting motor threshold (RMT) and self-reported fatigue (FSS-7) in the entire cohort of stroke patients (A), patients with right hemisphere strokes (B) and patients with left hemisphere strokes (C). FSS-7 was a significant predictor of RMT in patients with left hemisphere strokes.

Results of k-mean cluster analysis indicated that a 4-cluster solution produced the best-fit model, based on the highest silhouette score (0.311). The other models with 2, 3 and 5 clusters had a silhouette score of 0.283, 0.287 and 0.258 respectively. The number of objects in each cluster was 18, 14, 6 and 9. See supplementary methods for object-wise silhouette scores, distance from centroid and correlation with centroid. Cluster-wise data is shown in table 2. Kruskal-Wallis ANOVA was significant for 3 of the 4 factors tested (side of stroke H = 46, p<0.0001; HADS-D H=18.02, p<0.0001; HADS-A H=30.048, p<0.0001). Clusters 1 and 3 consisted of individuals with only left hemisphere stroke, and 2 and 4 consisted of only right hemisphere strokes. Post-hoc pairwise comparisons show a significant difference between clusters 1 and 3 (p<0.001), 1 and 4 (p=0), 2 and 3 (p=0), 2 and 4 (p=0.002) in both Depression and Anxiety scores, all of which survived a Bonferroni correction. Motor function PCA was different between clusters 1 and 2 (p=0.015) which did not survive Bonferroni correction for multiple comparisons. Figure 2 shows cluster-wise of Motor, HADS-depression, HADS-anxiety scores and FSS-7 scores.

**Table 2.** Cluster-wise mean (SD) values are displayed for all continuous variables and number of patients are displayed for categorical variables (Hemisphere affected, gender). P-value indicate the significance of Dunn’s post-hoc comparison, with cluster numbers within parenthesis showing which clusters the p value corresponds to.

|  | Cluster 1 (n=18) | Cluster 2 (n=14) | Cluster 3 (n=6) | Cluster 4 (n=9) | P-value |
| --- | --- | --- | --- | --- | --- |
| **FSS-7 Mean (SD)** | 5.45 (0.73) | 5.39 (1.00) | 5.3 (0.89) | 5.31 (0.59) | *NS* |
| **Hemisphere Affected** |  |  |  |  |  |
| Left \| Right | 18 \| 0 | 0 \| 14 | 6 \| 0 | 0 \| 9 | ***p* = 0.000** (1&2; 2&3; 1&4; 3&4) |
| **Gender** |  |  |  |  |  |
| Females \| Males | 7 \| 11 | 5 \| 9 | 4 \| 2 | 6 \| 3 | *NS* |
| **Age**<sub>years</sub> | 61.27 (11.53) | 57.07 (12.27) | 62.33 (16.66) | 54.44 (11.74) | *NS* |
| **Grip**<sub>% unaffected hand</sub> | 91.82 (24.79) | 84.66 (17.24) | 93.98 (9.82) | 94.3 (37.05) | *NS* |
| **NHPT**<sub>% unaffected hand</sub> | 99.28 (29.52) | 76.54 (21.62) | 91.66 (22.04) | 85.39 (18.54) | *NS* |
| **HADS**<sub>Depression</sub> | 4.94 (2.89) | 3.78 (1.31) | 8.0 (3.26) | 8.88 (1.52) | ***p* < 0.001** (1&3, 1&4, 2&3, 2&4) |
| **HADS**<sub>Anxiety</sub> | 3.61 (2.05) | 4.28 (1.62) | 12.66 (2.42) | 9.22 (1.93) | ***p* < 0.001** (1&3, 1&4, 2&3, 2&4) |
| **RMT**<sub>Affected Hemisphere (% MSO)</sub> | 52.15 (13.64) | 43 (11.02) | 43.5 (7.58) | 47.33 (10.42) | *NS* |
| **Motor PCA**<sub>AvgF1,F2 Grip, NHPT, RMT</sub> | 0.35 (0.78) | -0.37 (0.6) | 0.03 (0.59) | -0.14 (0.89) | *p* = 0.015 (1&2) |

**Figure 2.**
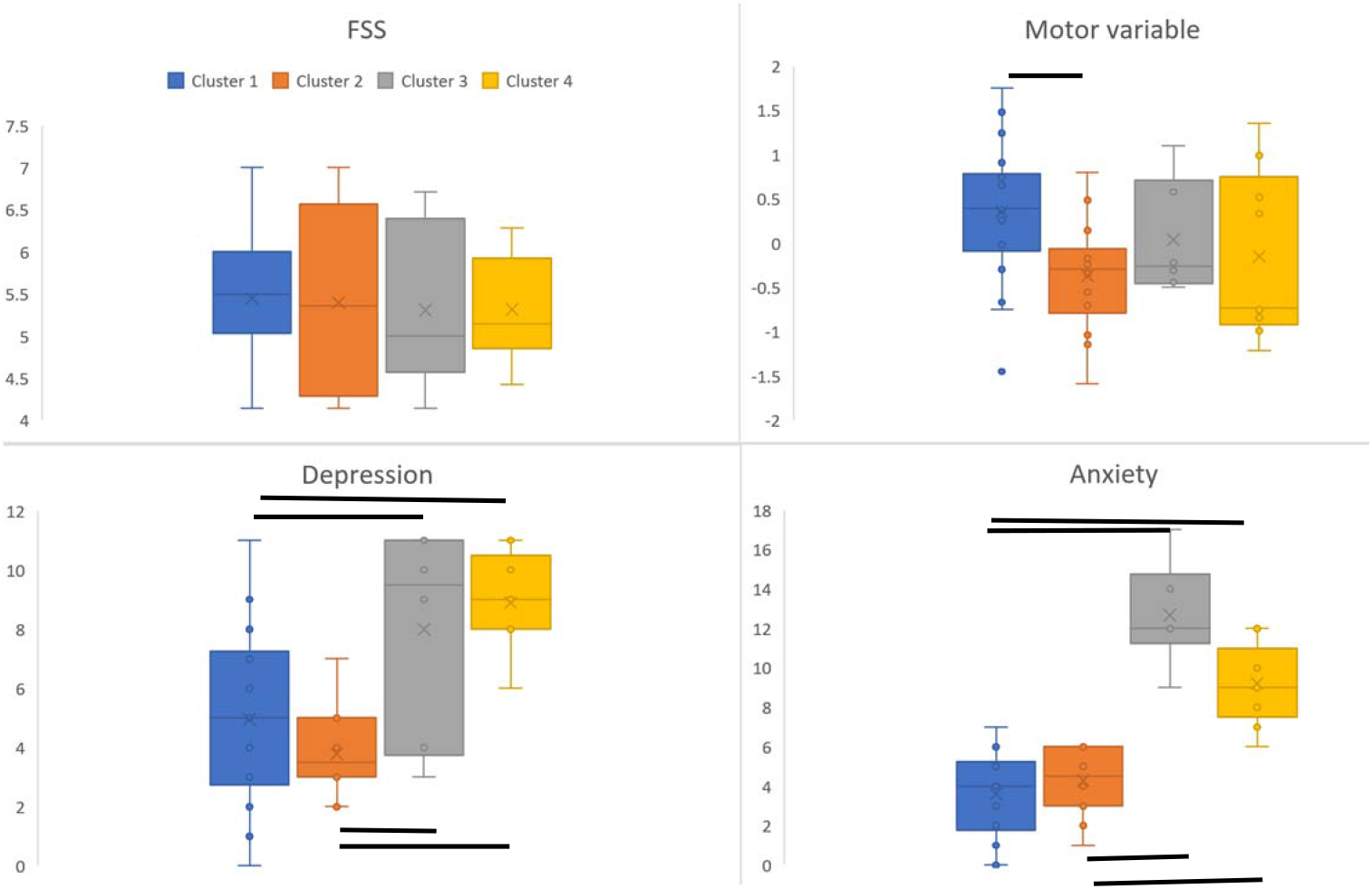
Box and whisker plots showing differences between clusters in the motor variable, depressive symptoms and anxiety. Clusters 1 and 3 are left hemispheric strokes while clusters 2 and 4 are right hemispheric strokes. Black lines represent significant differences between clusters.

## Discussion

The main finding in this study is the emergence of 4 distinct phenotypes within high fatigue chronic stroke survivors, using an unsupervised clustering method that included variables of motor behaviour, motor neurophysiology, side of stroke and scores of depression and anxiety. Of the four clusters, two were composed of only left hemispheric stroke and two right hemispheric stroke. In left-hemispheric strokes, the two clusters were differentiated by high and low levels of depressive symptoms and anxiety. Similarly, in right hemispheric strokes, levels of depressive symptoms and anxiety differentiated the two groups. There was a difference between left and right hemispheric non-depressive, non-anxious clusters, in the motor variable. While gender differences across the groups did not reach significance, there were more males in the non-depressive, non-anxious clusters with more females in the more anxious depressive clusters. In the entire sample of chronic stroke survivors, there was a significant positive relationship between fatigue levels and motor function, depression, anxiety, with only left hemisphere stroke displaying a positive correlation between fatigue and corticospinal excitability.

The emergence of 4 distinguishable clusters in mild to moderately impaired chronic stroke survivors, shows for the first time, it is possible to categorise high fatigue into smaller non-overlapping groups. In other disorders which present with complex behavioural profiles, such as autism spectrum disorder^19^ and psychosis^20^, phenotyping has been useful in identifying underlying pathophysiology and developing phenotype specific interventions. Here we propose phenotyping post-stroke fatigue serves a similar purpose by helping define the characteristics of high fatigue groups, and is in line with recommendations for post-stroke fatigue research^21^.

The difference in hemisphere affected, accompanied by differences in function and mood in the different clusters of high fatigue is a novel finding in this study. The relationship between post-stroke fatigue, impairment and mood have been previously investigated. However, the results have been confusing with some showing no relationship between fatigue and impairment^22,23^, while others revealing more nuanced interactions, such as, controlling for gender allowed for significant relationships with impairment and mood to emerge^10^. In some studies where impairment was not included, mood appeared to be the main driver of post-stroke fatigue^5^. While most previous investigations have included stroke hemisphere as a factor of interest, they all failed to reveal a role of stroke side in fatigue, possibly because both hemisphere strokes are equally likely to develop high fatigue, as shown by equal overall numbers of high fatigue in both right and left hemisphere strokes in this study. However, what has been revealed here are the factors essential for side of stroke to influence development of fatigue in an individual. Several such examples of how the current data might explain discrepancies in the literature can be elaborated, but, larger studies with more variables are necessary before definitive conclusions about high fatigue clusters are made. High levels of depressive symptoms and anxiety appear to be equally present in both right and left hemisphere strokes, although fewer individuals in both categories present this profile.

There are no clear differences seen in motor threshold that distinguishes left and right hemisphere strokes with high fatigue, despite the overall sample showing a positive relationship between threshold and fatigue levels in left hemisphere strokes. Corticospinal excitability (CSE) assessed using TMS thresholds is highly variable in the general population^24^, evident from the threshold values of the entire cohort in the current study. CSE of the left and right hemispheres behave differently during motor control with asymmetries in CSE seen between the two hemispheres^25–27^. The left hemisphere plays a dominant role in motor control characterized by higher CSE when compared to the right hemisphere, broader activation patterns during movement and greater thickens of the motor cortex^26,28,29^. The asymmetry in CSE between the two hemispheres is driven by inter-hemispheric connectivity. In a normal brain, inter-hemispheric connectivity is characterized by a net left-to-right inhibitory dominance and right-to-left excitatory dominance within the motor cortices and outside primary motor areas^30–33^, resulting in higher CSE of the left hemisphere when compared to the right. In post-stroke fatigue, opposite patterns of inter-hemispheric connectivity in primary motor cortices is seen with a shift from net left-to-right to a net right-to-left inhibitory dominance^34^. Given the shift in inter-hemispheric inhibition balance, those with high fatigue and left hemisphere stroke will present with lower excitability than those without fatigue. Results of the current study are in line with this expectation, however, the opposite is expected in right hemisphere strokes i.e. higher excitability associated with high fatigue. Despite a trend, this did not reach significance in the current study.

### Limitations

A noteworthy limitation of this study is the small numbers used to perform clustering analysis. The use of k-means clustering method was chosen over other more sophisticated clustering methods such as Latent Class Clustering and Gaussian Mixture Models due to the small number of datapoints, as k-means is known to perform better with smaller datasets. Moreover, by carefully selecting minimum number of variables with known associations with fatigue, the robustness of procedures has been maintained. Several previous studies have shown significant associations with cognitive dysfunction^7,35–37^. The focus of this paper was mainly physical parameters, and inclusion of cognitive measures could result in more clusters. The exclusion of cognitive measures can be seen as a limitation.

## Conclusion

Clustering revealed distinct sub-categories of high post-stroke fatigue defined by side of stroke, impairment and mood. Such clustering of high fatigue is essential to understand how fatigue emerges in a given population and is critical for developing effective interventions. The demonstration of capability of clustering methods to inform phenotyping of post-stroke fatigue is a significant step forward and a major contribution of this paper. Building on these foundations, future investigations must focus on definitively establishing phenotypes of post-stroke fatigue in larger samples with carefully selected structural, physiological, behavioural and self-reported variables associated with post-stroke fatigue.

## Data Availability

Data available on request

## Acknowledgments

We thank Mr Cameron Cook and the Clinical Research Network for their help with recruitment. We thank our lab manager Mr Paul Hammond for the technical support throughout the project. We extend our heartfelt thanks to all our stroke survivor participants in this study without whose enthusiasm and commitment this study would not have been possible.

## Funding

This work was supported by the Wellcome Trust (202346/Z/16/Z).

## Conflict of Interest

The authors report no conflict of interest.

## Notes

### Competing Interest Statement

The authors have declared no competing interest.

### Author Declarations

London Bromley Research Ethics Committee (REC reference number: 16/LO/0714)

